# Risk of SARS-CoV-2 exposure among hospital healthcare workers in relation to patient contact and type of care

**DOI:** 10.1101/2021.01.28.21250664

**Authors:** Susanna Klevebro, Fuad Bahram, K. Miriam Elfström, Ulrika Hellberg, Sophia Hober, Simon Kebede Merid, Inger Kull, Peter Nilsson, Per Tornvall, Gang Wang, Kalle Conneryd Lundgren, Sari Ponzer, Joakim Dillner, Erik Melén

## Abstract

**Aim:** We aimed to assess the risk for severe acute respiratory syndrome coronavirus-2 (SARS-CoV-2) infection in a large cohort of healthcare workers (HCWs).

**Methods:** From May 11 until June 11, 2020, 3,981 HCWs at a large Swedish Emergency Care hospital provided serum samples and questionnaire data. Exposure was measured by assaying IgG antibodies to SARS-CoV-2.

**Results:** The total seroprevalence was 17.7% and increased during the study period. Among the seropositive HCWs, 10.5% had been entirely asymptomatic. Participants who worked with COVID-19 patients had higher odds for seropositivity: ORadj 1.96 (95% CI 1.59 – 2.42). HCWs from three of the departments managing COVID-19 patients had significantly higher seroprevalences, whereas the prevalence among HCWs from the Intensive Care Unit (also managing COVID-19 patients) was significantly lower.

**Conclusion:** HCWs in contact with SARS-CoV-2 infected patients had a variable, but on average higher, likelihood for SARS-CoV-2 infections.

## Background

In February 27, the first confirmed case of SARS-CoV-2 infection in Stockholm, Sweden was diagnosed.^1^ The number of patients in need of hospital care due to COVID-19 in the Stockholm region escalated in March and the beginning of April and started to decline in May.^2^ Healthcare workers (HCWs) with direct patient contact are more exposed to the virus and thereby have a higher risk of contracting SARS-CoV-2 compared to the general population.^3–5^ Presence of SARS-CoV-2 IgG antibodies indicates a previous SARS-CoV-2 infection. A global review of previous studies published up to August 2020 found an overall seroprevalence of SARS-CoV-2 antibodies among HCWs to be 8.7% (95% CI: 6.7 – 10.9%).^6^ The aim of this study was to assess the seroprevalence in a large cohort of HCWs at a Swedish emergency care hospital after the first wave of COVID-19 patient admissions.

## Methods

Stockholm South General Hospital (Södersjukhuset) has one of the largest Emergency Care Departments in Northern Europe with about 550 in-patient beds. The hospital provides both general and specialized acute as well elective care. From February 27, when the first case was verified in the region, until June 30, 1,384 patients with confirmed SARS-CoV-2 infection had been admitted to the hospital (online supplement). Guidelines recommended the use of face shields, face masks and plastic aprons during direct contact with patients suspected to be infected. Aerosol filtering face masks were recommended during aerosol generating procedures, and in the ICU. HCWs were advised to stay at home if symptomatic and until feeling well for at least two days but were not routinely tested for SARS-CoV-2 during this period.

This cross-sectional study was initiated as a part of a regional study of HCWs conducted in the major emergency care hospitals in Stockholm, and all HCWs were invited to participate.

Stockholm South General Hospital had 4,641 employees in May 2020 and an estimate of the participation rate in this study is therefore 3981/4641 (85.8%). Blood samples were collected from May 11 to June 11, 2020. Self-reported information regarding current workplace, active work with infected patients (suspected or confirmed COVID-19 cases), and self-suspicion of prior SARS-CoV-2 infection were collected at the time of sampling.

Serum samples were prepared from whole blood, inactivated at 56° C for 30 minutes and stored at -20° C until analysis. IgG reactivity was measured towards three virus protein variants; Spike trimers comprising the prefusion-stabilized spike glycoprotein ectodomain,^7^ Spike S1 domain, and Nucleocapsid protein using multiplex bead array. Evaluation of the serology assay demonstrated a 99.2% sensitivity and 99.8% specificity.^3^

Results are presented as prevalence and odds ratio (OR) with 95% confidence intervals (CI). Age, sex, date of test, active work with COVID-19 patients, and department were included in the multivariable logistic regression model. Statistical analyses were performed in R.^8^ The study was approved by the Stockholm Ethical Review Board (dnr 2020-0162 and 2020-02724). All participants provided written informed consent and serology testing results were conveyed to participants.

## Results

In total, 3981 HCWs participated in the study. Participants had a mean age of 45.4 (11.9) years, 80.6% were female, and 64.3% responded that they had worked with patients with confirmed or suspected COVID-19.

Among the participating HCWs, 704 (17.7%) were seropositive for IgG antibodies against SARS-CoV-2. The prevalence increased during the test period (Figure 1) and was 19.5% among men and 17.3% among women (p=0.28). Among HCWs below the age of 40 years the prevalence was 20.0%, compared to 16.4% in HCWs 40 years of age and above (p=0.29).

**Figure 1:**
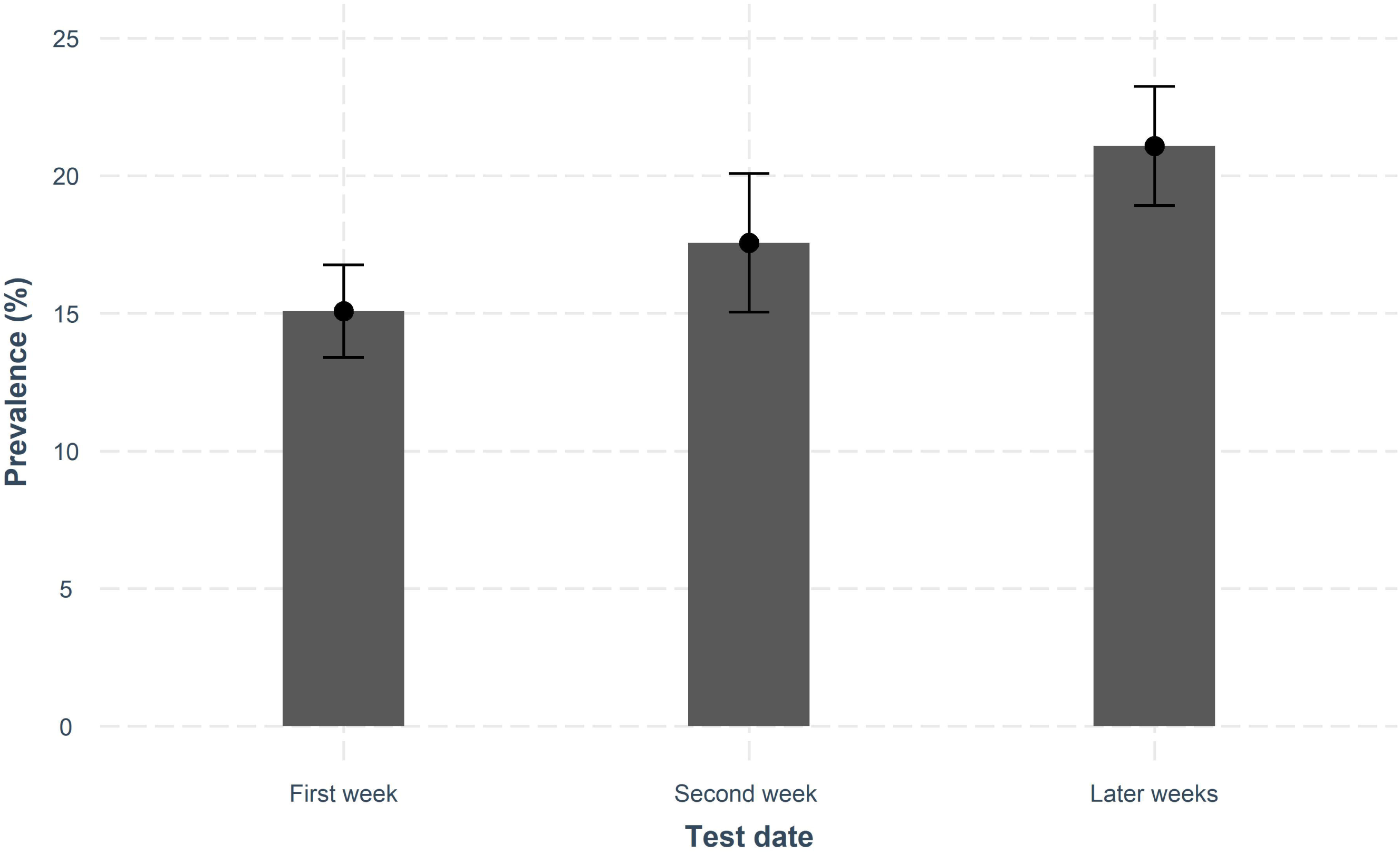
Prevalence of SARS-CoV-2 IgG antibodies by test date. Divided into First week, n=1,738; Second week, n=877; Later weeks (week 3 – 5), n=1,366 Error bars shows 95% confidence intervals.

HCWs who reported that they had worked with COVID-19 patients had a higher prevalence of antibodies, 21.1% compared to 11.7% without exposure, ORadj 1.96 (95% CI 1.59 – 2.42). Self-reported suspicion of COVID-19 symptoms prior to the study was associated with higher prevalence of a positive test, 46.4% compared to 5.5% among HCWs who had not suspected previous infection, (p<0.01). Among the seropositive HCWs, 10.5% had been asymptomatic (no suspicion of a previous SARS-CoV-2 infection).

Seroprevalence differed between departments: HCWs within the Departments of Cardiology, Infectious diseases, and Internal Medicine had a significantly higher prevalence compared to HCWs at the rest of the hospital after adjustment for sex, age, date of test, and active work with COVID-19 patients (Figure 2). In contrast, HCWs from the Departments of Anesthesiology/ ICU, Obstetrics/ Gynecology, and Pediatrics had a lower prevalence compared to the rest of the hospital.

**Figure 2:**
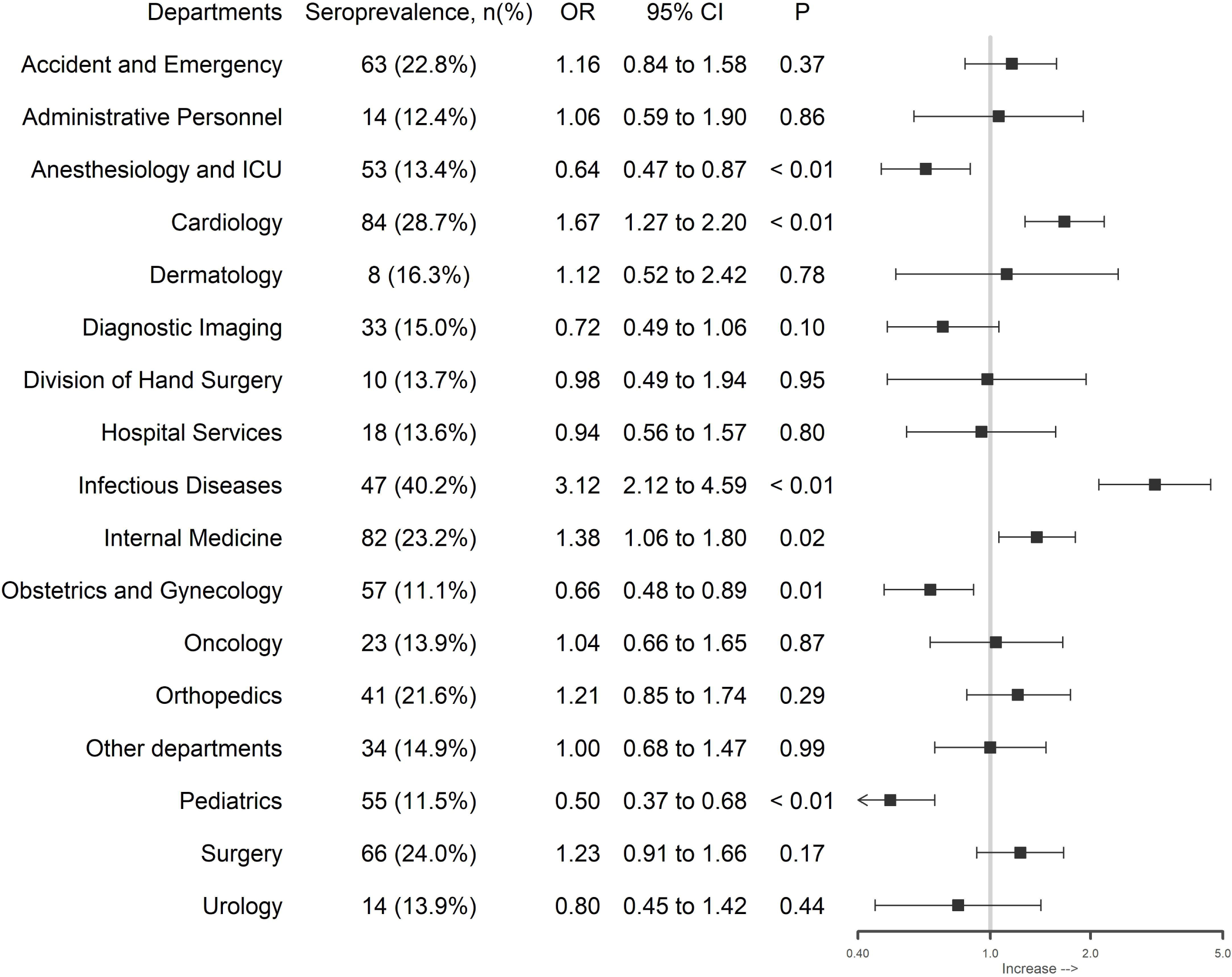
Differences between departments. The seroprevalence within the department is presented in parentheses, and the figure demonstrates Odds ratio (OR) and 95% confidence intervals (CI) from multivariable regression models comparing the department with all other departments adjusted for sex, age, date of test, and active work with COVID-19 patients.

## Discussion

This study found a prevalence of 17.7% of IgG antibodies against SARS-CoV-2 among HCWs in a large cohort of HCWs from Stockholm, Sweden sampled during the later phases of the first wave of the epidemic. The seroprevalence was similar to the 19.1% reported among 2,149 HCWs at Danderyd Hospital, a hospital of equivalent size in the Stockholm area,^3^ whereas Karolinska University Hospital demonstrated a lower seroprevalence of 11.5% among 12,928 HCWs.^9^ Globally, seroprevalences among HCWs have ranged from 0% to 45.3%.^6^ Residents in Stockholm had a seroprevalence of around 10% during the time of our study.^10^ Our results indicate that HCWs caring for COVID-19 patients had an increased risk of infection with SARS-CoV-2 during the first wave of the pandemic, but that this risk varied greatly.

Self-reported high suspicion of infection has previously been associated with seropositivity.^11–13^ In our study, 10.5% of the seropositive HCWs did not suspect a previous SARS-CoV-2 infection and were thus likely to have had an asymptomatic infection. The proportion of infected individuals that have asymptomatic infections is somewhat variable in the literature, with a recent systematic review estimating about 30%.^14^

As in previous studies,^3–5^ an increased prevalence was seen in HCWs who had been in contact with COVID-19 patients. Lower prevalence in the Pediatrics and Obstetrics/ Gynecology Departments was to be expected due to low exposure to patients infected with SARS-CoV-2 in these settings. HCWs from the Departments of Cardiology, Infectious diseases, and Internal Medicine were highly involved in the care of COVID-19 patients. The higher seroprevalence in these departments indicates an increased risk of infection for HCWs in these settings, which could not be explained by age, sex, patient contact or date of test.

Interestingly, as in the study by Grant et al.,^4^ HCWs at Anesthesiology/ ICU had a lower seroprevalence compared to the rest of the hospital. This could reflect the use of enhanced personal protective equipment or that the COVID-19 patients at the ICU may be in a later stage of the disease when viral loads of patients might have decreased.

During the spring, many HCWs were relocated within the hospital. Current workplace was self-reported, and a limitation of this study is that we were not able to fully distinguish primary (regular) workplace from workplace during the period of interest. Furthermore, we did not collect information regarding use of protective equipment or other potential exposures such as contact with infected persons outside work or with infected HCWs at work.

The number of COVID-19 patients admitted to our hospital peaked in the weeks before this study. The fact that the seroprevalences increased with calendar time suggests that the first wave of the epidemic was still ongoing during the time of this study. However, PCR testing for the virus was not performed.

## Conclusion

The prevalence of SARS-Cov-2 IgG antibodies was higher in HCWs caring for COVID-19 patients. However, the risk was highly variable between departments suggesting that other factors are important for the risk of the infection.

## Data Availability

Please contact Prof. E Melen with data requests.

## Acknowledgements

We would like to thank all HCWs at Södersjukhuset for participating in the study, and all the staff at Södersjukhuset and Karolinska University Hospital directly involved in the study. We are thankful to the SciLifeLab Autoimmunity and Serology profiling facility and all associated personnel for performing the serology analysis. We would also like to thank Jonas Blomqvist, Karolinska University Hospital, for study support.

## Tables

**Table 1:** Adjusted ORs for a positive SARS-CoV-2 IgG test in multivariable analyses

| Variables | OR | 95% CI | P |
| --- | --- | --- | --- |
| Age, Higher or equal than 40 years<sup>1</sup>, n=2,506 | 1.10 | 0.92 - 1.30 | 0.29 |
| Sex, Male<sup>2</sup>, n=771 | 1.12 | 0.91 - 1.37 | 0.28 |
| Test date, second week<sup>3</sup>, n=877 | 1.16 | 0.93 - 1.44 | 0.19 |
| Test date, third to fifth week<sup>3</sup>, n=1,366 | 1.49 | 1.24 - 1.80 | < 0.01 |
| Active work with COVID-19 patients<br>Yes<sup>4</sup> n=2,560 | 1.96 | 1.59 - 2.42 | < 0.01 |
| Active work with COVID-19 patients<br>Do not know<sup>4</sup> n=283 | 1.03 | 0.69 - 1.55 | 0.88 |

<sup>1</sup> Reference group: Age < 40 years, n=1,475
<sup>2</sup> Reference group: Female, n=3,210
<sup>3</sup> Reference group: Test date, first week, n=1,738
<sup>4</sup> Reference group: Active work with COVID-19 patients, no n=1,122
OR Odds Ratio; CI Confidence Interval; P p-value

