## Supplementary material for "Risk of SARS-CoV-2 exposure among hospital healthcare workers in relation to patient contact and type of care": online supplement

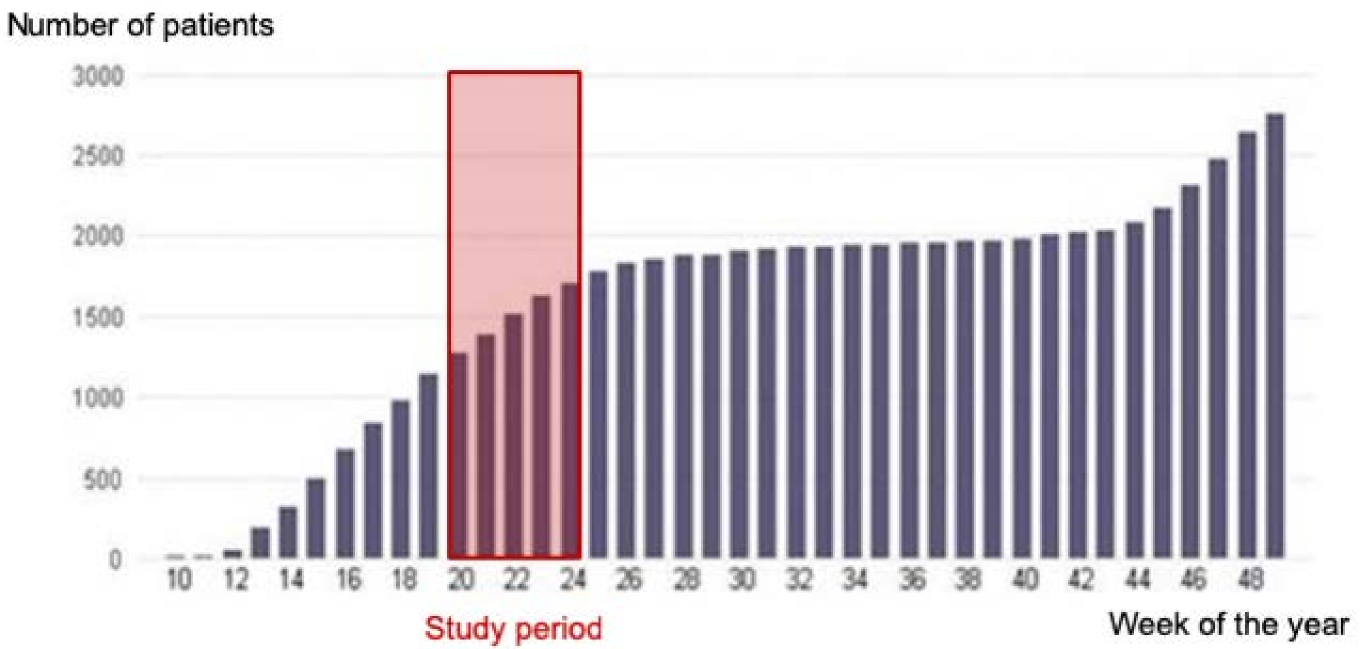


Cumulative number of patients with confirmed SARS-CoV-2 infection admitted to Stockholm South General Hospital (Södersjukhuset) 2020
